# Performance of an evidence-based risk algorithm to diagnose chlamydia and gonorrhea among pregnant Rwandan women

**DOI:** 10.64898/2026.05.18.26353484

**Authors:** Tyronza Sharkey, Julien Nyombayire, Rachel Parker, Rosine Ingabire, Claudine Umuhoza, Jean Bizimana, Jeannine Mukamuyango, Marie Aimee Unyuzimana, Amelia Mazzei, Amanda Tichacek, Susan Allen, Etienne Karita, Kristin M. Wall

## Abstract

Reproductive tract infections (RTI) are associated with adverse outcomes in pregnant African women. However, many diagnostic strategies are unaffordable or perform poorly. Here, we assess RTI prevalence and predictors of chlamydia/gonorrhea (CT/NG) in pregnant women reporting vaginal discharge and the performance of our previously published CT/NG risk algorithm in this population versus Rwandan National Guidelines (RNG). From 2017-2020, free sexually transmitted infections (STI) services were provided to residents in Kigali, Rwanda. Medical history and gynecologic examination were done. Laboratory assessments included HIV; syphilis; microscopy for trichomoniasis, bacterial vaginosis (BV), and candida; and PCR for CT/NG. Eighty-seven pregnant women received STI services. Prevalence was 28% for CT/NG, 15% for trichomoniasis, 24% for BV, 39% for candida, and 79% for any RTI. Predictors of CT/NG were age <=25 (adjusted prevalence odds ratio [aPOR]=4.92; 95% confidence interval [CI]: 1.52-15.90; p=0.008), inconsistent condom use (aPOR=4.86; 95%CI: 0.98-24.10; p=0.053), absence of candida (aPOR=4.23; 95%CI: 1.13-15.82; p=0.032), and endocervical inflammation/discharge (aPOR=4.91; 95%CI: 1.40-17.20; p=0.013). Our algorithm outperformed the 2019 and 2024 RNG (sensitivity: 92% versus 46% and 35% respectively). Pregnant women seeking STI services had high RTI prevalence. Our algorithm performed well. Algorithms tailored for pregnant women and including male partner risk factors should be explored.

## INTRODUCTION

Ineffective sexually transmitted infection (STI) prevention, diagnosis, and treatment efforts have led to global resurgence of STI, with pregnant women in low and middle income countries being disproportionately affected [1]. Increasing rates of chlamydia (CT), gonorrhea (NG), and trichomoniasis (TV) in African pregnant women are alarming given their associations with preterm birth, low birth weight, small for gestational age, perinatal mortality, stillbirth, and maternal and mother-to-child HIV risk [2, 3]. Bacterial vaginosis (BV) and vaginal candidiasis (VCA) are common and associated with miscarriage, preterm birth, and low birth weight [3-5]. Improved diagnosis and treatment of reproductive tract infections (RTI, herein referring to NG, CT, TV, BV, and VCA) common during pregnancy are urgently needed to reduce unnecessarily high rates of adverse pregnancy outcomes.

In many African countries, including Rwanda, etiologic tests for these RTI are not routine largely due to cost and logistics. Rather, syndromic management is the norm. Per the 2019 Rwanda National Guidelines (RNG) risk algorithm [6], women presenting at public health clinics with vaginal discharge (regardless of pregnancy) must meet ≥2 of the following 3 risk factors to receive treatment for CT/NG: age <21, single, ≥2 sexual partners in the last 30 days. While a first step towards a feasible public health approach to diagnosing CT/NG, this algorithm is uninformed by local epidemiology. More recent RNG (2024) recommend use of a gynecologic exam to guide treatment for CT/NG [7]. Per the current RNG, patients with vaginal discharge are treated for CT/NG if they have either: 1) lower abdominal or cervix pain found on gynecologic exam (bimanual and speculum) and no delayed/absent monthly periods, no recent abortion, no vaginal bleeding, no pelvic mass or tenderness and/or 2) muco-purulent discharge from cervix or cervicitis but no lower abdominal or cervix pain on bimanual exam [7]. However, gynecologic exams may not be feasible in many Rwandan health centers due to availability of trained staff and equipment [8]. Additionally, since many RTI are asymptomatic, syndromic approaches fail to detect many cases [9].

Additionally, there are a dearth of non-laboratory RTI diagnostic approaches tailored for pregnant women, creating a significant public health gap in the prevention of preterm birth and other negative pregnancy outcomes. Importantly, risk factors for RTI are quite different for pregnant African women as most are married and have a single partner. Finally, as vaginal discharge is normal in pregnancy, syndromic approaches may lead to overtreatment [10].

We previously developed and evaluated a risk algorithm for CT/NG using locally derived epidemiological variables for symptomatic, primarily non-pregnant women in Kigali [11, 12]. Our CT/NG algorithm had a sensitivity of 79%-91%, outperforming the 2019 RNG which had a sensitivity of 26% [11]. We also found that microscopic diagnosis of TV, BV, and VCA at our site was feasible and low-cost.

Here, we determine RTI prevalence among pregnant women in Kigali seeking care at an STI clinic, identify their risk factors for CT/NG, and assess the performance of our previously published algorithm, which was not derived in a pregnant population, to identify CT/NG in pregnant women.

## METHODS

### Aims

Our study aims to assess the prevalence of RTI and CT/NG co-infections in Rwandan pregnant women seeking STI services. In addition, our study aims to expand the application of our previously published CT/NG risk algorithm, not derived in a pregnant population, to pregnant women and compare its performance against the current and prior Rwandan National Guidelines.

### Ethics

STI services were anonymous, free of charge, and carried out in accordance with Rwandan and US government guidelines and regulations. Provision of STI prevention and treatment services was determined to be non-research by the Emory University Institutional Review Board (IRB) and Rwandan National Ethics Committee (RNEC). Thus, written informed consent was waived by the RNEC, Emory IRB, and the US Centers for Disease Control funded by PEPFAR.

### Setting

Between March 2017 and November 2020, clients sought free STI services at the Center for Family Health Research (CFHR) STI clinic in Kigali, Rwanda.

### Study design

Our cross-sectional study highlights women, particularly pregnant women, who sought free STI testing and treatment services at the CFHR STI clinic. Though some clients were seen at subsequent visits for retesting, repeat/subsequent infections, or additional treatment, we report only the initial STI clinic visit here.

### Client recruitment

CFHR used radio announcements to encourage Kigali residents with genital discharge, ulcer, or discomfort complaints to seek free STI services at CFHR clinic. Referral slips from clients who received STI services and from pharmacists providing treatment for suspected STI were also used to recruit. CFHR clients diagnosed with an STI could opt-out of nurse-facilitated partner notification [6, 7].

### Data collection and diagnostic procedures

As part of STI services, medical history (demographic, sexual behavior, and STI symptoms) and gynecologic examination were done. Medical history data were collected using standardized nurse-administered surveys. Demographic data collected included age, living/marital status, educational level, employment status, and pregnancy status. Sexual behavioral data collected included number of sexual partners, condom use, number of children under 18 years in the household, and number of additional children desired. Self-reported symptom data collected included genital itching, burning sensations when passing urine, genital ulcer, dyspareunia, unpleasant odor, and lower abdominal pain. This survey was informed by literature review and our previous STI work in Rwanda [13-15] including Rwandan HIV programs. Gynecologic exams and endocervical/vaginal swab collection were performed by CFHR research nurses. As part of gynecologic exams, clinical findings such as external vaginal inflammation and discharge, internal vaginal inflammation and discharge, endocervical inflammation or discharge, and genital ulcers were reported. CFHR laboratory staff performed HIV rapid testing; rapid plasma reagin for syphilis; and vaginal wet mount microscopy to diagnose TV, BV, and VCA. Endocervical swabs were tested using GeneXpert (Cepheid) PCR to diagnose NG and CT. Laboratory results and participant responses to surveys were entered into Microsoft Access. RTI were treated in accordance with the RNG [6, 16].

### Risk algorithm development

The derivation, calibration, and internal and external validation of our CT/NG risk algorithm, not derived in a pregnant population, have been previously published [11]. We chose a vaginal discharge-based algorithm to align with the RNG [6, 7]. Of note, vaginal discharge has been cited as the most common STI symptom among Rwandan women [12, 17]. Briefly, we used standard methods including logistic regression models to identify factors associated with CT/NG infection among a derivation cohort of n=468 women (8.5% of whom were pregnant) generating weighted score values by dividing each variable’s estimated model coefficient by the smallest coefficient and rounding to the nearest integer. The Hosmer-Lemeshow Goodness-of-Fit test assessed model calibration, and 10-fold cross validation methods [18] were used for internal validation. The risk algorithm included being ≤25 years of age, having no/primary education, sometimes using condoms, and not reporting genital itching each worth 2 points while not having full-time employment, not having candida, and having BV were each worth 1 point. The risk algorithm had a sensitivity of 91% and a specificity of 36% at a score cut-off of >=4 [11].

### Data analyses

The present analysis was a subset of women receiving STI services who self-reported pregnancy and vaginal discharge. The distribution of RTI, HIV, syphilis, co-infections with CT/NG in all women, and CT/NG risk factors in pregnant women only were described using descriptive statistics (counts and percentages for categorical variables and medians and interquartile ranges for continuous variables).

Composite variables were created for gynecologic exam findings. Vaginal inflammation and vaginal discharge were combined as discharge is a common symptom of vaginitis. An either/or ‘endocervical inflammation or discharge’ composite was created as presence of either indicates possible cervicitis. Treatment indications of suspected vaginitis or cervicitis are key components of RNG.

We identified demographic, sexual behavioral, symptom, laboratory, and gynecologic factors associated with CT/NG using Chi-square, Fisher’s exact, or Mann-Whitney U tests, as appropriate. Variables associated with CT/NG infection in bivariate analyses (p<0.1) were assessed for multi-collinearity; if any variables were found to be collinear, the variable with the strongest association with the outcome was considered for multivariable modelling. Multivariable logistic regression models were then built with backwards elimination, and adjusted prevalence odds ratios (aPORs), 95% confidence intervals (CI), and corresponding p-values were reported. Missing data were summarized in tables.

We also assessed the performance of our previously published CT/NG algorithm, which was not derived in a pregnant cohort, at different score cut-offs to identify CT/NG in pregnant women. Score cut-offs represent the median score and one point above and below the median.

Sensitivity, specificity, positive predictive value (PPV), and negative predictive value (NPV), and area under the receiver operating characteristic curve (AUC) were calculated for our CT/NG algorithm and compared the analogous performance characteristics for the 2019 and 2024 RNG CT/NG algorithms. To compare RNG 2024, we used self-reported as a proxy for clinical finding of lower abdominal pain. Data were analysed in SAS 9.4 (Cary, NC).

## RESULTS

### Prevalence of RTI, HIV, syphilis, and co-infections with CT/NG in women with self-reported vaginal discharge by pregnancy status (Table 1)

To assess RTI prevalence, we described and tabled the distribution of RTI, HIV, syphilis and co-infections in Rwandan women by pregnancy status. Of the 1,355 women receiving STI services and self-reporting vaginal discharge, eighty-seven (6%) were pregnant. Most pregnant women (79%) had at least one RTI. The prevalence of NG was 20% and CT was 18% in pregnant women. The prevalence of CT and NG infection was 28%. The prevalence of other RTI was 39% for VCA, 24% for BV, and 15% for TV in pregnant women. Ten percent of pregnant women were co-infected with CT and NG. Other prevalent co-infections with CT and/or NG (>=5%) in pregnant women were CT and TV; NG and BV; CT, NG, and TV; and CT, NG, and VCA.

**Table 1.** Distribution of RTI, HIV, Syphilis and co-infections among pregnant and non-pregnant women with vaginal discharge ^a^.

|  | Pregnant<br>(N=87) |  | Non-pregnant<br>(N=1268) |  |
| --- | --- | --- | --- | --- |
|  | n | Col% | n | Col% |
| No RTI <sup>b</sup> | 17 | (20) | 360 | (29) |
| Any RTI (CT/NG/TV/BV/VCA) <sup>b</sup> | 69 | (79) | 843 | (67) |
| Any RTI, HIV and syphilis <sup>b</sup> | 70 | (80) | 890 | (71) |
| NG | 17 | (20) | 245 | (19) |
| CT | 16 | (18) | 200 | (16) |
| TV <sup>c</sup> | 13 | (15) | 124 | (10) |
| VCA <sup>c</sup> | 34 | (39) | 283 | (23) |
| BV <sup>c</sup> | 21 | (24) | 361 | (29) |
| HIV <sup>c</sup> | 6 | (7) | 146 | (12) |
| Syphilis <sup>c</sup> | 5 | (6) | 83 | (7) |
| **Co-infections with CT and/or NG** |  |  |  |  |
| CT or NG | 24 | (28) | 371 | (29) |
| CT and NG | 9 | (10) | 74 | (6) |
| CT and TV | 7 | (8) | 30 | (2) |
| NG and BV | 6 | (7) | 93 | (7) |
| CT, NG, and TV | 4 | (5) | 11 | (1) |
| CT, NG, and VCA | 4 | (5) | 4 | (0) |
| CT and syphilis | 3 | (4) | 21 | (2) |
| NG and syphilis | 3 | (4) | 28 | (2) |
| CT and BV | 2 | (2) | 89 | (7) |
| NG and HIV | 2 | (2) | 53 | (4) |

<sup>a</sup> Co-infections with less than 3% prevalence in non-pregnant and pregnant women are not tabled.
<sup>b</sup> Women who were missing any laboratory result and had no positive laboratory results were excluded from calculations of 'No RTI,' 'Any RTI,' or 'Any RTI, HIV and syphilis' composites (n=18).
<sup>c</sup> Among non-pregnant women, missing data were noted for TV (n=13), BV (n=17), VCA (n=15), syphilis (n=25), and HIV (n=19). Among pregnant women, missing data were noted for syphilis (n=2). Abbreviations: RTI, reproductive tract infection; HIV, human immunodeficiency virus; STI, sexually transmitted infection; CFHR, Center for Family Health Research in Rwanda; CT, chlamydia; NG, gonorrhea; TV, trichomoniasis; BV, bacterial vaginosis; VCA, vaginal candidiasis; col, column.

For comparison, in 1,268 non-pregnant women seen at CFHR during the same timeframe, the prevalence of NG, CT, VCA, and TV were lower than pregnant women while BV, HIV, and syphilis prevalence was higher. In addition, fewer non-pregnant women were co-infected with CT and NG. In non-pregnant women, prevalent co-infections (>=5%) included CT and NG; CT and BV; and NG and BV. Most non-pregnant women had at least one RTI.

### Demographic and sexual behavior characteristics of pregnant women with self-reported vaginal discharge by CT/NG status (Table 2a)

To identify risk factors for CT/NG in pregnant women, we first summarized the distribution and performed bivariate analysis of demographic and sexual behavior characteristics of pregnant women by CT/NG status in Table 2a. Our bivariate analysis found that pregnant women with CT/NG were more likely to be younger (<=25 years of age), less likely to be married/cohabiting, more likely to report having >1 partner in the last 30 days, more likely to report using condoms sometimes versus never using condoms/no sex in the last three months, and reporting having fewer children under 18. These characteristics differed statistically (p <0.1) by CT/NG status and will be included in the multivariate analysis. Characteristics that did not differ based on chlamydia/gonorrhea status included referrer for STI services, educational level, employment status, and not tabled, number of days since sexual contact you suspected RTI was acquired from.

**Table 2a.** Distribution of demographic and sexual behavioral characteristics and bivariate factors associated with CT/NG in pregnant women with self-reported vaginal discharge.

|  | Total<br>(N=87) |  | Either CT or NG<br>(n=24) |  | CT and NG<br>Uninfected (n=63) |  |  |
| --- | --- | --- | --- | --- | --- | --- | --- |
|  | n/<br>median | Col%/<br>IQR | n/<br>median | Col%/<br>IQR | n/<br>median | Col%/<br>IQR | p-value |
| Demographics |  |  |  |  |  |  |  |
| Age (years) | 27.0 | 8.0 | 20.5 | 7.0 | 28.0 | 8.0 | <0.001 |
| Age |  |  |  |  |  |  |  |
| 25 or younger | 40 | (46) | 18 | (75) | 22 | (35) | 0.001 |
| Older than 25 | 47 | (54) | 6 | (25) | 41 | (65) |  |
| Referrer |  |  |  |  |  |  |  |
| Heard from Friends/Walk-in | 59 | (66) | 19 | (79) | 40 | (63) | 0.162 |
| Radio Advert | 28 | (31) | 5 | (21) | 21 | (33) |  |
| Other<sup>a</sup> | 2 | (2) | 0 | (0) | 2 | (3) |  |
| Living & Marital Status |  |  |  |  |  |  |  |
| Married and Cohabiting | 63 | (72) | 14 | (58) | 49 | (78) | 0.070 |
| Other | 24 | (28) | 10 | (42) | 14 | (22) |  |
| Education Level |  |  |  |  |  |  |  |
| None/Primary | 34 | (39) | 11 | (46) | 23 | (37) | 0.426 |
| Secondary/Higher | 53 | (61) | 13 | (54) | 40 | (63) |  |
| Employment Status |  |  |  |  |  |  |  |
| Full-time employment | 27 | (31) | 5 | (21) | 22 | (35) | 0.204 |
| Part-time/Student/ Jobless | 60 | (69) | 19 | (79) | 41 | (65) |  |
| Sexual Behaviors |  |  |  |  |  |  |  |
| No. partners in last 30 days<sup>b</sup> | 1.0 | 0.0 | 1.0 | 0.0 | 1.0 | 0.0 | 0.086 |
| No. partners in last 30 days |  |  |  |  |  |  |  |
| None or one partner | 76 | (88) | 18 | (78) | 58 | (92) | 0.123 |
| More than one partner | 10 | (12) | 5 | (22) | 5 | (8) |  |
| Condom use during vaginal sex in the last three months<sup>b</sup> |  |  |  |  |  |  |  |
| Sometimes | 12 | (14) | 6 | (26) | 6 | (10) | 0.076 |
| Never/No vaginal sex in the last three months | 74 | (86) | 17 | (74) | 57 | (90) |  |
| No. of children under 18 | 1.0 | 2.0 | 0.0 | 1.0 | 1.0 | 2.0 | 0.008 |
| No. of additional children desired | 1.0 | 1.0 | 1.5 | 1.0 | 1.0 | 1.0 | 0.956 |

<sup>a</sup> Pharmacy/Other/Index Partner Invitation.
<sup>b</sup> Missing data were noted for number of sexual partners, condom use, number of days since sexual contact you suspect RTI was acquired from (n=1).
Abbreviations: CT, chlamydia; NG, gonorrhea; col, column; IQR, interquartile range; RTI, reproductive tract infection.

### Symptom, laboratory, and gynecologic characteristics of pregnant women with self-reported vaginal discharge by CT/NG status (Table 2b)

Next, as part of identifying risk factors for CT/NG in pregnant women, we summarized the distribution and performed bivariate analysis of self-reported STI symptoms, laboratory, and gynecologic characteristics of pregnant women by CT/NG status in Table 2b. Our bivariate analysis showed that pregnant women with CT/NG were less likely to have VCA though more likely to have TV and syphilis. In addition, pregnant women with CT/NG infection were more likely to have endocervical inflammation or discharge findings on gynecologic exam. These characteristics differed statistically (p <0.1) by CT/NG status and will be included in the multivariate analysis.

Characteristics that did not differ based on chlamydia/gonorrhea status included self-reported symptoms, HIV and BV status, and gynecologic findings of external vaginal inflammation and discharge, internal vaginal inflammation and discharge, and genital ulcer.

**Table 2b.** Distribution of symptom, laboratory, and gynecologic characteristics and bivariate factors associated with CT/NG in pregnant women with self-reported vaginal discharge.

|  | Total<br>(N=87) |  | Either CT or NG<br>(n=24) |  | CT and NG<br>Uninfected<br>(n=63) |  |  |
| --- | --- | --- | --- | --- | --- | --- | --- |
|  | n/<br>median | Col%/<br>IQR | n/<br>median | Col%/<br>IQR | n/<br>median | Col%/<br>IQR | p-value |
| Self-reported symptoms<sup>a</sup> |  |  |  |  |  |  |  |
| Genital itching | 65 | (75) | 15 | (63) | 50 | (79) | 0.106 |
| Burning sensations when<br>passing urine | 40 | (46) | 12 | (50) | 28 | (44) | 0.642 |
| Genital ulcer | 7 | (8) | 3 | (13) | 4 | (7) | 0.386 |
| Dyspareunia<sup>b</sup> | 30 | (35) | 7 | (30) | 23 | (37) | 0.619 |
| Unpleasant odor<sup>b</sup> | 38 | (45) | 12 | (52) | 26 | (42) | 0.399 |
| Lower abdominal pain<sup>b</sup> | 33 | (39) | 10 | (43) | 23 | (37) | 0.592 |
| Days with symptoms<sup>b</sup> |  |  |  |  |  |  |  |
| 1-10 | 19 | (22) | 4 | (17) | 15 | (24) | 0.525 |
| 11 or more | 67 | (78) | 19 | (83) | 48 | (76) |  |
| HIV and other RTI/STI<br>Results <sup>a</sup> |  |  |  |  |  |  |  |
| HIV | 6 | (7) | 3 | (13) | 3 | (5) | 0.203 |
| RPR Result<sup>b</sup> | 5 | (6) | 4 | (17) | 1 | (2) | **0.021** |
| Trichomonas | 13 | (15) | 7 | (29) | 6 | (10) | **0.039** |
| Candida | 34 | (39) | 4 | (17) | 30 | (48) | **0.008** |
| Bacterial vaginosis | 21 | (24) | 7 | (29) | 14 | (22) | 0.499 |
| Gynecologic Exam<sup>c</sup> |  |  |  |  |  |  |  |
| External vaginal<br>inflammation and discharge | 66 | (75) | 16 | (70) | 50 | (77) | 0.341 |
| Internal vaginal inflammation<br>and discharge<sup>b</sup> | 62 | (77) | 16 | (70) | 46 | (79) | 0.351 |
| Endocervical inflammation or<br>discharge<sup>b</sup> | 37 | (44) | 15 | (65) | 22 | (35) | **0.014** |
| Genital ulcer<sup>b</sup> | 7 | (8) | 3 | (13) | 4 | (6) | 0.378 |

<sup>a</sup> For Yes/No responses for covariates related to self-reported STI symptoms and gynecologic exam, only 'Yes' responses were included on the table. For Positive/Negative responses for covariates related to HIV and other RTI/STI results, only 'Positive' results were included on the table.
<sup>b</sup> Missing data were noted for number of days with symptoms and gynecologic exam findings of external vaginal inflammation or discharge and genital ulcer (n=1); syphilis RPR result, self-reported dyspareunia, unpleasant odor, and lower abdominal pain, and gynecologic exam findings of endocervical inflammation or discharge (n=2); self-reported genital ulcer (n=3); and gynecologic exam findings of internal vaginal inflammation and discharge (n=6).
Abbreviations: CT, chlamydia; NG, gonorrhea; STI, sexually transmitted infection; col, column; IQR, interquartile range; RTI, reproductive tract infection; HIV, human immunodeficiency virus; RPR, syphilis rapid plasma reagin.

### Predictors of CT/NG infection in multivariate analysis (Table 3)

Next, we assessed all variables associated with CT/NG infection in the bivariate analysis (p<0.1) for multi-collinearity. No collinear variables were identified. In the final multivariable logistic regression model after backward elimination, age 25 or younger (adjusted prevalence odds ratio [aPOR]=4.92; 95% confidence interval [CI]: 1.52-15.90; p=0.008), report using condoms sometimes (aPOR=4.86; 95%CI: 0.98-24.10; p=0.053), not having candida (aPOR=4.23; 95%CI: 1.13-15.82; p=0.032), and presence of endocervical inflammation or discharge on gynecologic exam (aPOR=4.91; 95%CI: 1.40-17.20; p=0.013) were associated with CT/NG infection.

**Table 3.** Factors associated with CT/NG in pregnant women with vaginal discharge seeking STI services.

|  | Multivariable model |  |  |  |
| --- | --- | --- | --- | --- |
|  | aPOR | 95% CI |  | p-value |
|  |  | LL | UL |  |
| Demographics |  |  |  |  |
| Age, dichotomous |  |  |  |  |
| 25 or younger | 4.92 | 1.52 | 15.90 | 0.008 |
| Older than 25 | ref |  |  |  |
| Sexual behaviors |  |  |  |  |
| Condom use during vaginal sex in the last three months |  |  |  |  |
| Sometimes | 4.86 | 0.98 | 24.10 | 0.053 |
| Never/No vaginal sex in the last three months | ref |  |  |  |
| HIV and Other STI Results |  |  |  |  |
| Candida |  |  |  |  |
| Positive | ref |  |  |  |
| Negative | 4.23 | 1.13 | 15.82 | 0.032 |
| Gynecologic Exam |  |  |  |  |
| Endocervical Inflammation or Discharge |  |  |  |  |
| Yes | 4.91 | 1.40 | 17.20 | 0.013 |
| No | ref |  |  |  |

Abbreviations: CT, chlamydia; NG, gonorrhea; STI, sexually transmitted infection; CFHR, Center for Family Health Research in Rwanda; CI, confidence interval; aPOR, adjusted prevalence odds ratio; LL, lower limit; UL, upper limit; HIV, human immunodeficiency virus.

### Performance of the CT/NG risk algorithm in pregnant women (Table 4)

Each pregnant woman’s data for age, condom use, self-reported genital itching, candida diagnosis, educational level, employment status, and BV diagnosis was applied to the previously published algorithm’s risk factors, derived from a nonpregnant population, to diagnose CT/NG. The risk algorithm’s diagnosis was verified against the CT/NG PCR result. The CT/NG risk algorithm performed well in the pregnant cohort (AUC=0.78, 95%CI: 0.67, 0.88, p<0.001). The risk algorithm had 92% sensitivity, 51% specificity, 42% PPV, and 94% NPV for the median score cutoff of >=□4.

**Table 4.** Performance of CT/NG algorithm and RNGs to diagnose CT/NG in pregnant women with vaginal discharge.

|  | Sensitivity (%) | Specificity (%) | PPV (%) | NPV (%) |
| --- | --- | --- | --- | --- |
| **CFHR Algorithm<sup>a</sup>** |  |  |  |  |
| Score >= 3 | 92 | 30 | 33 | 90 |
| Score >= 4 | 92 | 51 | 42 | 94 |
| Score >= 5 | 75 | 59 | 41 | 86 |
| Score >= 6 | 58 | 81 | 54 | 84 |
| **2019 RNG<sup>b</sup>** |  |  |  |  |
| Score >=2 | 46 | 94 | 73 | 82 |
| Score = 3 | 8 | 97 | 50 | 73 |
| **2024 RNG<sup>c</sup>** |  |  |  |  |
| Treat for CT/NG (based on vaginal discharge symptom and gynecologic exam findings) | 35 | 76 | 36 | 75 |

<sup>a</sup> AUC=0.78, 95% CI: 0.67, 0.88, p<0.001.
<sup>b</sup> AUC=0.69, 95% CI: 0.56, 0.81, p=0.004.
<sup>c</sup> AUC = 0.56, 95% CI: 0.44, 0.67, p=0.340.
Abbreviations: CFHR, Center for Family Health Research in Rwanda; CT, chlamydia; NG, gonorrhea; RNG, Rwandan National Guidelines; STI, sexually transmitted infection; PPV, positive predictive value; NPV, negative predictive value; AUC, area under the curve.

### Performance of the RNG in pregnant women (Table 4)

Next, we applied each pregnant woman’s data for age, marital status, and number of sexual partners to the RNG 2019 and lower abdominal pain to the RNG 2024 to diagnose CT/NG. RNG diagnoses were verified against the CT/NG PCR result. Applying the 2019 RNG cutoff of >=*2 (the cutoff level for CT/NG treatment) to the pregnant cohort had a sensitivity of 46%, specificity of 94%, PPV of 73%, and NPV of 82%. The AUC for the 2019 RNG was also lower compared to the risk algorithm (AUC=0.69, 95%CI: 0.56, 0.81, p=0.004). Applying the new 2024 RNG for treatment of CT/NG to the pregnant cohort had a sensitivity of 35%, specificity of 76%, PPV of 36%, and NPV of 75%. Comparatively, the 2019 and 2024 RNG were less sensitive and had a lower PPV and NPV than the CFHR CT/NG risk algorithm.

## DISCUSSION

The high prevalence of curable RTI observed among this pregnant cohort is alarming as most presenting with vaginal discharge had at least one RTI. More than one-quarter of the pregnant cohort had either CT or NG. The pregnant cohort’s CT/NG prevalences were higher compared to pooled prevalence data from other pregnant women in Eastern Africa though similar for TV, BV, and VCA [19, 20]. A Rwandan cohort study in HIV and syphilis negative pregnant women, attending health centers in Kigali, found similar high CT rates (21.5%) at their study baseline though lower rates of TV, BV, and VCA (5.2%, 19.6%, and 21.5% respectively) [21]. Of note, studies compared were either exclusively asymptomatic [20] or did not indicate [19, 21]. Overall, these studies point to a high burden of RTI among pregnant women. In addition, the pregnant cohort had more RTI and CT and/or NG co-infections compared to non-pregnant women. Given this cohort’s high RTI burden and the deleterious effects of these infections during pregnancy, effective solutions are urgently needed to diagnose and treat RTI during pregnancy.

Risk factors for CT/NG in pregnant women included absence of candida, sometimes using condoms, age <=25 years old, and endocervical inflammation or discharge on gynecologic exam. A study in symptomatic Kenyan pregnant women found being younger (age <20 years old) as a risk factor for CT/NG but not condom use or clinical findings related to endocervical inflammation or discharge [22]. As our pregnant cohort is small, comparisons were not possible; thus, a larger study is needed. Endocervical inflammation or discharge on gynecologic exam was not included previously during our risk algorithm development in 2020 as routine gynecological exams were not considered feasible in public health centers at the time. Though both algorithms maintain vaginal discharge symptom as point of entry, the 2024 RNG no longer uses a risk factor algorithm but gynecologic exam findings to determine treatment for CT/NG. Though implementation of routine gynecologic exams in public health centers will require time, training, and equipment, our modeling data indicate that evaluation of endocervical inflammation or discharge may improve the performance of a CT/NG risk algorithm tailored for pregnant women.

Our risk algorithm for CT/NG outperformed the 2019 and 2024 RNG in pregnant women with sensitivity of 92% versus 46% and 35%, respectively. With our algorithm’s high sensitivity, pregnant women who need CT/NG treatment will be identified; however, the lower specificity (50% versus 94% and 76%, respectively) translates to instances of overtreatment due to false positives. RTI overtreatment is a growing concern due to increasing antimicrobial resistance, particularly for NG [16]. Given the morbidities or mortality associated with untreated RTI during pregnancy [2-5], and since no treatment contraindications for pregnancy have been cited in presumptive trials [23], our algorithm’s profile is still favorable. When deciding trade-offs between sensitivity and specificity, focus should generally be on high-sensitivity tests when infections have serious health consequences and the risks associated with overtreatment are acceptable [24].

Vaginal wet mount microscopy to diagnose TV, BV, and VCA is important for comprehensive RTI diagnoses and also supports our CT/NG algorithm. Though less accurate than molecular testing, microscopy has acceptable performance to diagnose TV, BV, and VCA when laboratory technicians are well trained [25]. In addition, microscopy can be performed in <1 hour allowing for same day treatment [25]. Furthermore, in resource limited settings vaginal swabs can be self-collected for microscopy, a more feasible option in clinic settings than speculum exam, which are not routinely done in practice. The only speculum exam covariate predictive of CT/NG in our study was the endocervical inflammation/discharge composite, observed in only 65% of the pregnant cohort, which was neither sensitive nor specific. As such, the feasibility of conducting routine vaginal wet mount microscopy at Rwandan public health centers requires further investigation. Public health center needs assessments conducted by CFHR suggest the availability of sufficient laboratory technicians and microscopes, though re/training for RTI diagnoses may be needed.

As the number of pregnant clients attending the CFHR STI clinic was relatively small, we were unable to derive a CT/NG algorithm specifically for pregnant women. We plan to collect risk factor data from a larger group of pregnant women to derive and validate a tailored algorithm for this population. Studies in Africa including ours have highlighted that risk factors for RTI during pregnancy may be different than those for non-pregnant populations, and importantly, consideration of men’s risk factors may greatly improve CT/NG algorithm accuracy in this population [26, 27]. Most pregnant women in this study were married and cohabiting. Thus, including male partner risk factors to improve diagnoses could be feasible in Rwanda as roughly 90% of men attend at least one antenatal clinic visit with their pregnant partner [15], presenting an opportunity to treat both partners together.

Our study had limitations. Our study findings are most generalizable to symptomatic pregnant women attending an urban public health facility, which is a population with a high prevalence of HIV/STI [17]. RTI risk factors in rural settings, among those who do not seek public health services, or among asymptomatic women may be different and should be explored. Our survey included limited assessment of social determinants of health indicators of RTI such as partner communication, stigma, empowerment, discrimination, and harassment [28] which could be explored for further algorithm refinement and to inform counseling and development of other RTI prevention interventions. As symptomatic pregnant women in this study sought STI services, the CT/NG prevalence may be overestimated. However, a few East African studies of mostly asymptomatic pregnant women have also reported similar high CT and/or NG prevalence (∼15% to 22%) [21, 29]. Risk factor data were self-reported thus introducing the possibility of recall and social desirability bias, which can result in underreporting risky behaviors; these sources of information bias may or may not be differentiated by CT/NG status. We did not routinely collect data on clinical finding of lower abdominal pain during gynecologic exam at CFHR, which is used in the 2024 RNG to decide treatment for CT/NG [7]. However, we did collect data on self-reported lower abdominal pain which was used as a proxy when evaluating 2024 RNG performance.

Overall, our study suggests a high prevalence of RTI and CT/NG coinfections in symptomatic pregnant women. We highlight that an evidence-based CT/NG risk algorithm, though not tailored for pregnant populations, outperformed RNG, which are based on World Health Organization (WHO) STI management recommendations [30]. Our findings also add to the literature on risk factors for CT/NG during pregnancy, which can be used to inform tailored algorithms. Finally, to our knowledge, no African country has adopted routine molecular testing for STI diagnosis due to resource constraints; however, as access to molecular testing increases in some locales, our inexpensive algorithm could be explored as screening tool for referral to molecular testing to increase the feasibility and affordability of molecular testing.

## CONCLUSION

Effective, sustainable methods to improve RTI diagnoses in pregnant women are needed to impact extreme rates of maternal and neonatal morbidity and death. Findings from this study may be applicable in public health facilities to improve RTI diagnoses in pregnant populations, and may motivate future work to develop improved, tailored algorithms for this high-need population.

## Data Availability

The datasets analyzed during the current study are available from the corresponding author on reasonable request and a signed data sharing agreement.

## Funding

This work was supported by the International AIDS Vaccine Initiative (IAVI) with the generous support of United States Agency for International Development (USAID). This work was partially funded by Center for Disease Control and Prevention (CDC); the National Institute of Allergy and Infectious Diseases at the National Institutes of Health [grant number R01 AI51231]; the AIDS International Training and Research Program Fogarty International Center [grant number D43 TW001042]; and the Emory Center for AIDS Research [grant number P30 AI050409]. The contents of this manuscript are the responsibility of the authors and do not necessarily reflect the views of IAVI, USAID, CDC, National Institutes of Health, Fogarty International Center, or the United States Government.

## Authors’ Contributions

TDS contributed to the interpretation of data, drafted the article and revised it critically for important intellectual content, and gave final approval of the version to be published. JN, RI, JB, JM, AM, MAU, AT, EK contributed to the conception and design of the study, reviewed the article critically for important intellectual content, and gave final approval of the version to be published. RP contributed to the analysis and interpretation of data, reviewed the article critically for important intellectual content, and gave final approval of the version to be published. SA contributed to the study design and conception, contributed to the analysis and interpretation of data, revised the article critically for important intellectual content, and gave final approval of the version to be published. KMW contributed to the analysis and interpretation of data, drafted the article and revised it critically for important intellectual content, and gave final approval of the version to be published.

## ADDITIONAL INFORMATION

### Competing Interests

The authors declare no competing interests.

## Notes

### Competing Interest Statement

Our manuscript is under review at a journal. We have no other competing interest to declare.

### Funding Statement

This study was funded by the Centers for Disease Control and Prevention, the National Institute of Allergy and Infectious Diseases (Grant ID R01 AI51231), the AIDS International Training and Research Program Fogarty International Center (Grant ID D43 TW001042), the Emory Center for AIDS Research (Grant ID P30AI050409), and IAVI.

### Author Declarations

Emory University Institutional Review Board waived ethical approval for this work; Rwandan National Ethics Committee waived ethical approval for this work; US Centers for Disease Control waived ethical approval for this work

